# Prevalence Of COVID-19 In Rural Versus Urban Areas in a Low-Income Country: Findings from a State-Wide Study in Karnataka, India

**DOI:** 10.1101/2020.11.02.20224782

**Authors:** Manoj Mohanan, Anup Malani, Kaushik Krishnan, Anu Acharya

## Abstract

Although the vast majority of confirmed cases of COVID-19 are in low- and middle-income countries, there are relatively few published studies on the epidemiology of SARS-CoV-2 in these countries. The few there are focus on disease prevalence in urban areas. We conducted state-wide surveillance for COVID-19, in both rural and urban areas of Karnataka between June 15-August 29, 2020. We tested for both viral RNA and antibodies targeting the receptor binding domain (RBD). Adjusted seroprevalence across Karnataka was 46.7% (95% CI: 43.3-50.0), including 44.1% (95% CI: 40.0-48.2) in rural and 53.8% (95% CI: 48.4-59.2) in urban areas. The proportion of those testing positive on RT-PCR, ranged from 1.5 to 7.7% in rural areas and 4.0 to 10.5% in urban areas, suggesting a rapidly growing epidemic. The relatively high prevalence in rural areas is consistent with the higher level of mobility measured in rural areas, perhaps because of agricultural activity. Overall seroprevalence in the state implies that by August at least 31.5 million residents had been infected by August, nearly an order of magnitude larger than confirmed cases.

## Introduction

There are few published studies on the epidemiology of SARS-CoV-2 in low- and middle-income countries, which contain the vast majority of confirmed cases. India has the second-highest number of reported cases, but most seroprevalence estimates have come from urban centers.

Urban areas, because of higher population densities, are thought to be more vulnerable to COVID-19. However, rural areas received millions of migrant workers fleeing cities and agriculture was an essential-activity exempt from lockdown.

We conducted surveillance for COVID, both viral RNA and antibodies targeting the receptor binding domain (RBD), on a representative population from urban and rural areas of the Indian state of Karnataka (population 64.06 million), from June 15-August 29, 2020.

## Methods

### Study Sample and Location

Our study sample was drawn from an existing, representative sample of a panel survey, the Center for Monitoring Indian Economy’s Consumer Pyramids Household Survey (CPHS). The CPHS, collected by the Center for Monitoring Indian Economy (CMIE), is the world’s largest longitudinal household panel data set. It includes roughly 174,00 households (and their roughly 1.2 million members) nationwide. The CPHS has been surveying these household 3 times annually since 2014 and includes a range of questions on household financial and social status.

### Details on CPHS sampling frame

The CPHS survey has a stratified, multiple-stage design. The primary sampling units (PSU’s) are villages and towns defined by the 2011 Indian Census. The ultimate sampling units (USU’s) are the households from these PSUs.

#### Selection of Primary Sampling Units

The broadest stratum for the survey is a “homogenous region” (HR), an areas comprised of neighboring districts in a state where the districts have similar agro-climactic conditions, similar urbanization rates, and similar female literacy. Table 1 provides the districts in the 5 CPHS homogenous regions in Karnataka.

**Table 1.**
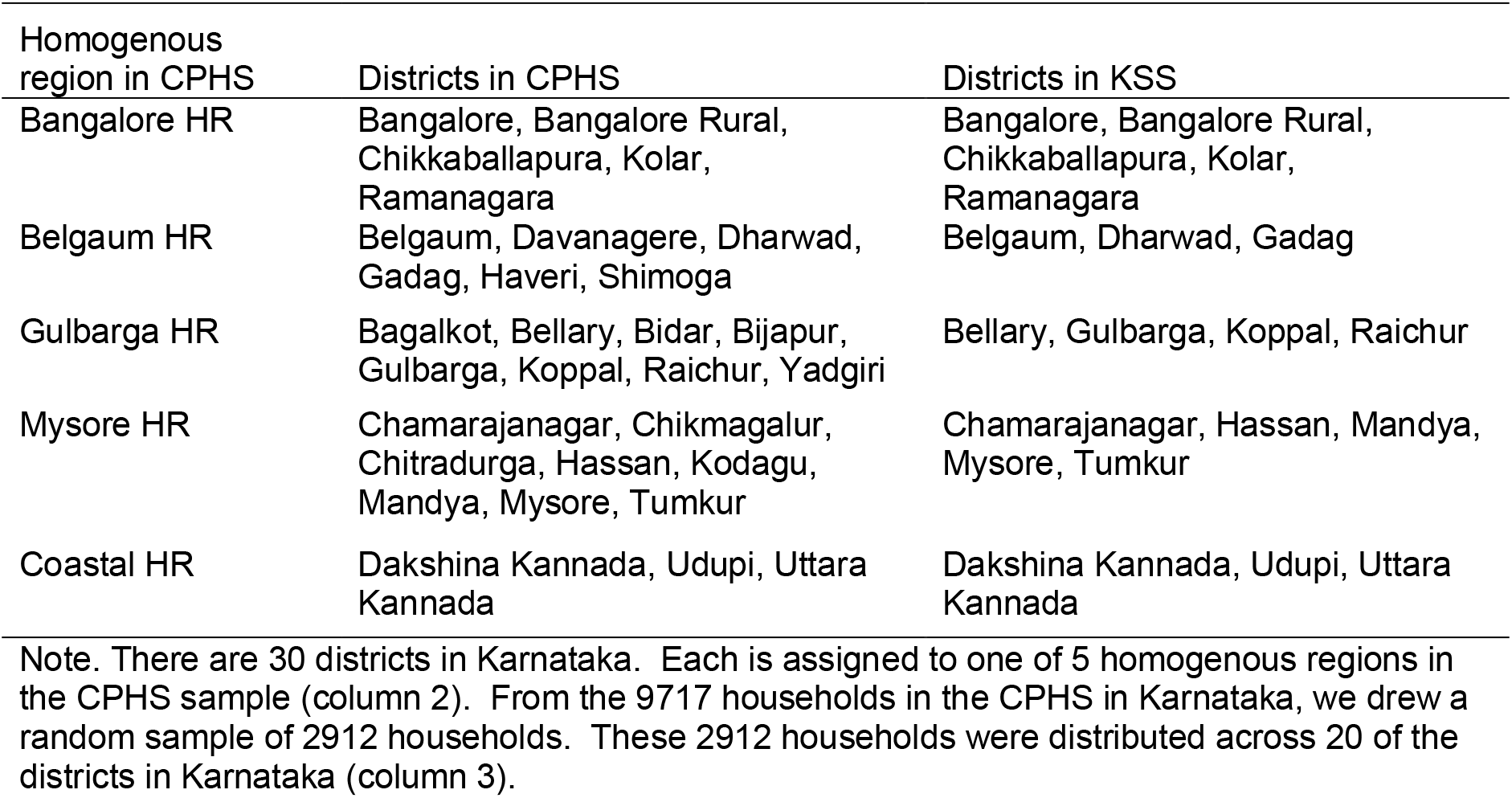
**The districts of Karnataka included in each homogenous region in the CMIE Consumer Pyramids Database (CPHS) and in the Karnataka Seroprevalence Survey (KSS) sample.**

| Homogenous region in CPHS | Districts in CPHS | Districts in KSS |
| --- | --- | --- |
| Bangalore HR | Bangalore, Bangalore Rural, Chikkaballapura, Kolar, Ramanagara | Bangalore, Bangalore Rural, Chikkaballapura, Kolar, Ramanagara |
| Belgaum HR | Belgaum, Davanagere, Dharwad, Gadag, Haveri, Shimoga | Belgaum, Dharwad, Gadag |
| Gulbarga HR | Bagalkot, Bellary, Bidar, Bijapur, Gulbarga, Koppal, Raichur, Yadgiri | Bellary, Gulbarga, Koppal, Raichur |
| Mysore HR | Chamarajanagar, Chikmagalur, Chitradurga, Hassan, Kodagu, Mandya, Mysore, Tumkur | Chamarajanagar, Hassan, Mandya, Mysore, Tumkur |
| Coastal HR | Dakshina Kannada, Udupi, Uttara Kannada | Dakshina Kannada, Udupi, Uttara Kannada |

Note. There are 30 districts in Karnataka. Each is assigned to one of 5 homogenous regions in the CPHS sample (column 2). From the 9717 households in the CPHS in Karnataka, we drew a random sample of 2912 households. These 2912 households were distributed across 20 of the districts in Karnataka (column 3).

Within each HR, CPHS samples from 2 strata: urban and rural areas. Within urban areas, which are the towns from the 2011 Indian Census, CPHS samples from 4 strata defined by town size. Within each town stratum, CPHS selects at least one town via simple random selection. Within the rural strata, CPHS picks a random subset of villages via simple random selection. In Karnataka, CPHS has picked 31 – 51 villages per HR and 3-4 towns per HR.

#### Ultimate sampling units

Towns are divided by into Census Enumeration Blocks, a cluster of 100-125 neighboring households. In towns selected for the urban strata, CPHS chooses a random subsample of the town’s CEBs via simple random selection. A minimum of 21 CEBs are chosen per town. Within each CEB, it conducts systematic random sampling of every nth households, where n was a randomly chosen integer from 5 to 15. Within a CEB, CPHS aims to select 16 households.

In rural strata villages, a different approach was used. Households were selected by, first, selecting the central street and, then, conducting systemic random sampling as in urban strata towns. Within a village, CPHS aims to select 16 households.

Overall, the CPHS sample frame includes 9717 households that reside in Karnataka, a state with a population of roughly 64 million persons.

### Sample selection for the Karnataka Seroprevalence Study

Our study, which we label the Karnataka Seroprevalence Study (KSS), draws a random sample from CPHS’s 9717 households in Karnataka separately for the urban and rural strata. In the urban strata towns, we selected on quarter of the CEBs selected for the CPHS sample, rounding up to the nearest integer. Within each CEB, we attempted to survey all households in the CPHS sample.

To select from villages, we made one accommodation to logistics. We limited the sample to villages within 30 km of the centroid of each town in our KSS sample. This restriction was critical because there was a lockdown in effect during our sampling and finishing quickly was critical as seroprevalence changes over time. With each CPHS village that met this criterion, we attempted to survey all CPHS households in that village.

There were some cases where we were not able to execute the survey in a CEB or village because of the lockdown restrictions and imposition of containment areas. Before going to that CEB in a town or that village, we replaced it in the sample. In the case of CEBs, we randomly selected another CEB from the CPHS sample in the same town. In the case of villages, we selected the nearest villages to the inaccessible village.

Although there are 30 districts in Karnataka, our random selection of one-quarter of towns or villages from the CPHS sample of CEBs resulted in administrative units not being selected in 10 districts. Hence our study sample includes 20 districts.

### Sample size and minimum detectable effect

We did not conduct power calculations when selecting our sample. First, our main constraint was how many households from the CPHS frame we were allowed to include in our serological study. The owner of the database, CMIE, was concerned that requesting biosamples might cause their panel households to refuse to participate in CPHS going forward. Weighing the value of this study against that risk, they were only willing to sacrifice roughly one-quarter of the sample.

Second, we did not have strong priors on the fraction of the population that had previously been exposed. The fraction of the population that previously had a confirmed case was small when we started the study on June 15, 2020. That fraction was an implausible estimate of positive proportion on an ELISA test. A more realistic higher number would require a larger sample size to estimate given the standard sample size formula for binary outcomes.

However, we were able to calculate the minimum detectable effect (MDE) given the sample size we were allocated. This is summarized for different priors estimates of positive proportions in Figure 1. Our calculation of MDE comes from the usual sample size formula for a two-sided test and binary outcomes:

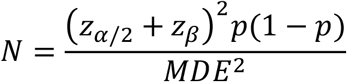

where one seeks 95% confidence (*Z*_*α/2*_ *=* 1.96) and 80% power (*Z*_*β*_ *=* 0.84), *p* is a prior estimate of positive proportion, and *N* is the sample size.

**Figure 1.**
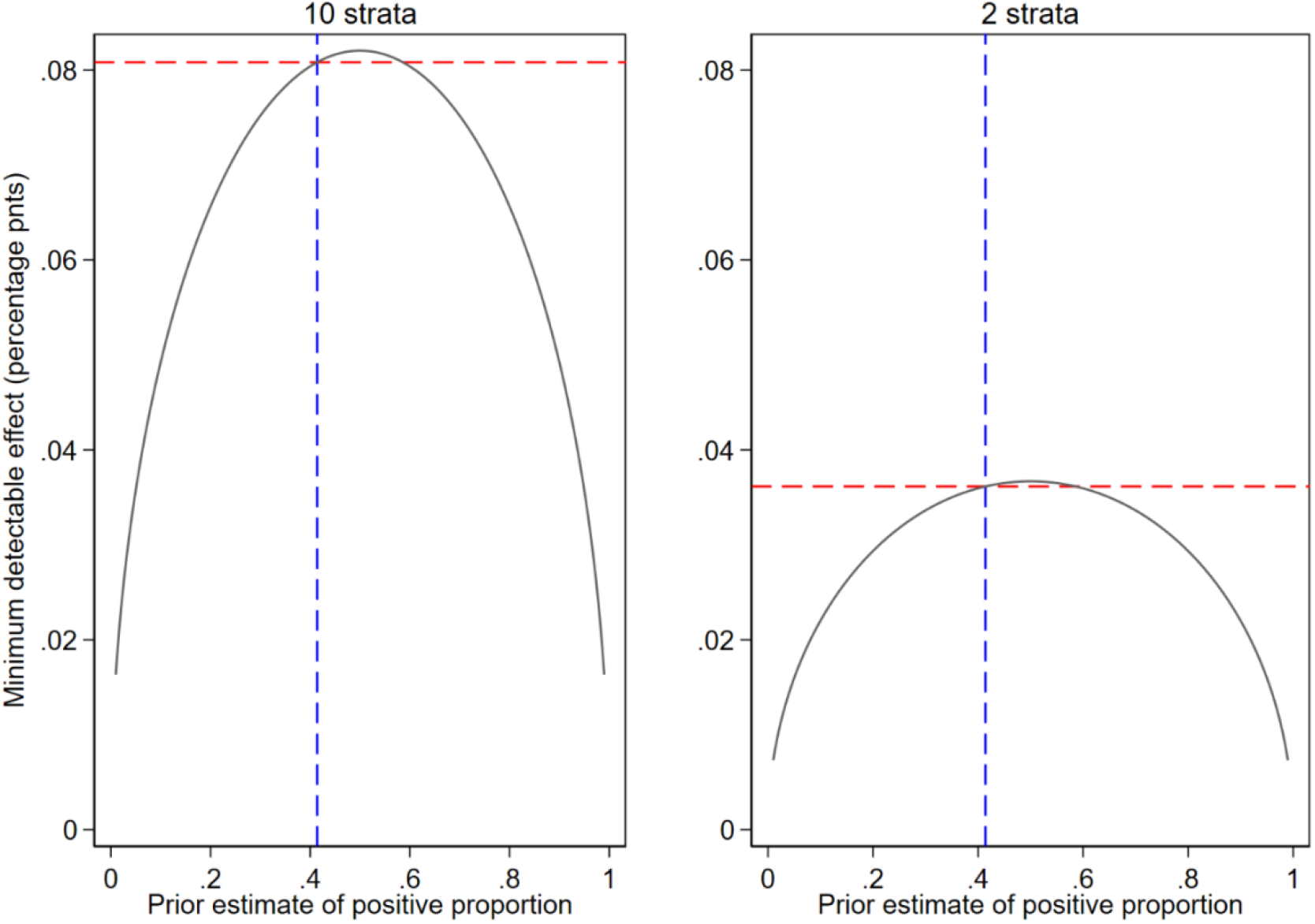
Minimum detectable effect for different priors, by number of strata in the study. Note. The dashed blue line shows the actual state-wide positive proportion estimated from the data (41.4%). The dashed red line shows the minimum detectable effect (MDE) if the prior on positive proportion were equal to the estimate state-wide positive proportion. The panel on the left in Figure 1 shows what the MDE would be if the study examined 10 strata (5 homogenous regions x urban or rural areas) or 2 strata (urban versus rural areas).

The dashed blue line in the shows the actual state-wide positive proportion estimated from the data (41.4%). The dashed red line shows the minimum detectable effect (MDE) if the prior on positive proportion were equal to the estimate state-wide positive proportion. The subplot on the left shows what the MDE would be if the study examined 10 strata (5 homogenous regions x urban or rural areas) or 2 strata (urban versus rural areas). In the former (latter) case, if the prior were fortuitously the state-wide proportion, the MDE would be 8.08 (3.61) percentage points.

We ultimately chose to estimate positive proportion in 10 strata, which implies our MDE would have been 8.08 percentage points if (a) our belief was that there was no difference between positive proportion in urban and rural areas and (b) our prior estimate for positive proportion in each strata turned out to be the correct estimate for the state.

### Study duration

Because prevalence changes over time, we estimate prevalence in each stratum (homogenous region x urban status) within a 2-3 week window. Karnataka is the 6th largest Indian state by area (191,791 km2). The data collection was conducted by the study team in various parts of the state from June 15 to August 29, 2020, to complete sampling across the entire state. The median date on which we visited each strata is depicted on the x-axis in the Figures 4 and 5 in the results section.

### Data collection and Consent

At each of the 2912 households in the KSS sample, we first sought consent from anyone at home to complete a health survey that asked about demographics, comorbidities, travel and contact history, and COVID symptoms. Multiple individuals at each household were allowed to complete this survey.

At each of the households wherein at least 1 person consented to complete the health survey, we asked 1 person to consent to a 5ml blood draw (via venipuncture using an EDTA vacutainer) and a nasopharyngeal nasal swab. The blood was refrigerated until it was delivered to Xcyton lab. The swab was placed in viral transport medium (VTM) and refrigerated until delivered to Aster Labs. The VTM we used was the Covisafe™ kit (manufactured by Mapmygenome) for collecting oropharyngeal swabs that make it possible to transport the swabs at ambient temperatures even if refrigeration failed.

### Attrition

Before visiting each household, we endeavored to call the household to increase the probability that we would arrive when they were at home. Despite this effort, we lost 580 households (20.0% of 2912 households in the KSS sample frame) because they were not home when we arrived (Figure 2).

**Figure 2.**
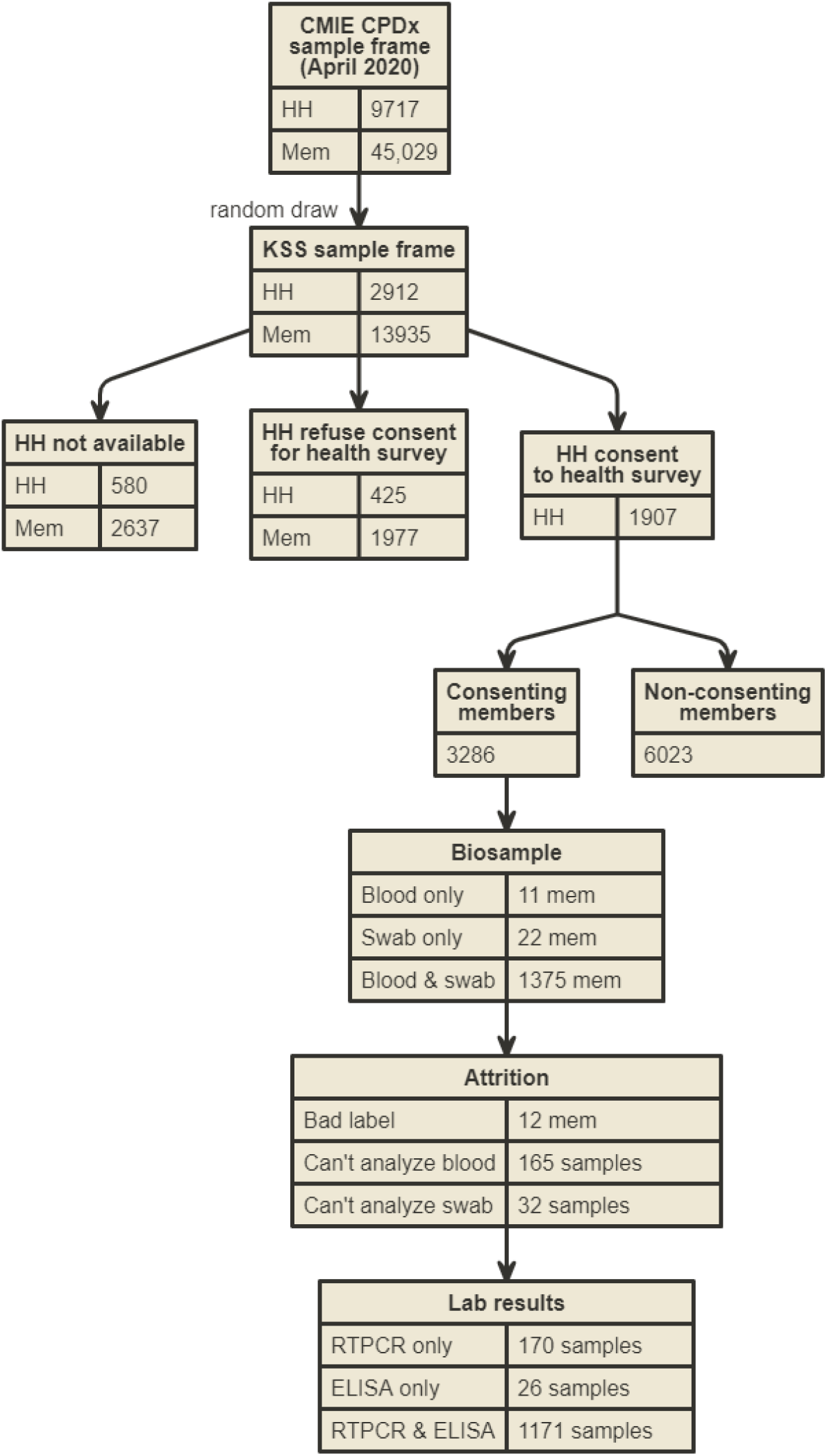
Flow chart of study sample.

When visiting a household, we sought consent to conduct our health survey. 425 households (14.6% of all 2912 households in the KSS frame or 18.2% of 2332 available households) declined consent to participate. In total 1907 households had at least 1 person consent to complete a health survey (demographic and health questionnaire). This implies a response rate of 65.5% of the 2912 households in the KSS frame and 81.8% of households that were available. We note that these consent rates are comparable to other studies of COVID.^1^

Across the households that consented to the health survey, there were 1363 persons who consented to provide both blood and swab. In 2 households one person provided blood and another provided a swab. In 11 (22) households, the oner person who consented to a biosample only provided person blood (swab). In total, 1374 households (72.1% of the 1907 households where someone consented to a health survey) had a person that provided blood and 1385 households (72.6% of 1907) had a member that provided a swab.

We were unable to obtain lab results for all samples. Among 1374 blood samples, we were unable to obtain results for 170 samples (12.3% of blood samples)). Of these, 12 were lost because labels were unreadable by the lab and 158 had inadequate blood drawn to extract serum. Among swab samples. We were unable to obtain law results for 46 samples (3.3% of swab samples). Of these 12 were due to bad labels and the result were due to failure in the VTM. (See Table 2 for details on sample composition by consent and lab specimen availability, by location and date)

**Table 2.**
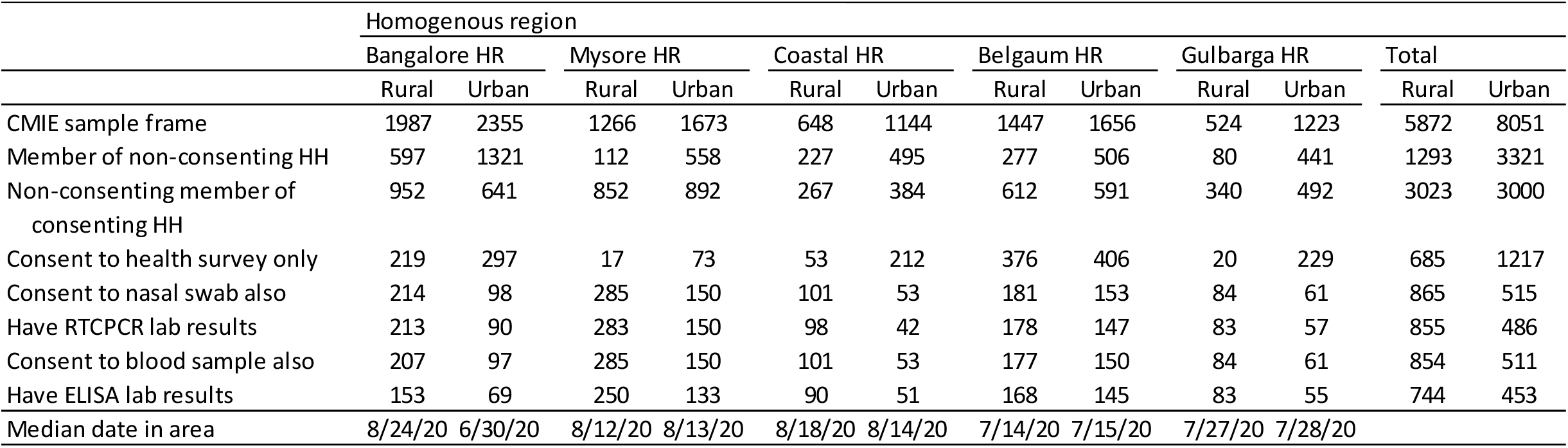
Sample composition by consent status and lab results, by location and date.

### Testing

At Xcyton Labs, plasma was separated and tested for IgG antibodies to the receptor binding domain (RBD) of the SARS-CoV-2 virus using an ELISA test developed by Translational Health Science and Technology Institute, India. This ELISA test is positive if IgG score, defined as the ratio of the titer in a sample and in a negative control, is greater than 1.5. This test has sensitivity of 84.7% (95% CI: 80.6–88.1%) and specificity of 100% (95% CI: 97.4-100)^2^.

Aster Labs conducted RTPCR tests targeting the N gene using the ARGENE® SARS-COV-2 R-GENE® assay from Biomerieux SA^1^. This test received Emergency Use Authorization from the FDA in May 2020^2^. We code borderline results on this test are as negative results. This test has sensitivity of 100% (95% CI: 87.7-100%) and specificity of 100% (98.1-100%)^3^.

### Outcomes

Our primary outcome is the proportion of positive results on ELISA tests for each of 10 homogenous region x urban/rural strata. Our secondary outcomes are:

- Adjusted proportion of positive results on RT-PCR tests for each of 10 homogenous region x urban/rural strata,
- Adjusted proportion of positive results on ELISA tests for the 5 homogenous regions,
- Adjusted proportion of positive results on ELISA tests for urban or rural areas,
- Adjusted seroprevalence based on the ELISA test accounting for the imperfect accuracy of those tests.

### Statistical methods

We estimated the adjusted proportion of positive tests 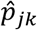 people in community type *j ∈* (urban, rural) in homogenous region *k* as a weighted average of test results for all individuals who are in our sample with test results and reside in strata (*j, k*). The weight for a strata is the normalized product of two factors: (a) a weights to ensure that the members of our sample in a strata are presentative of the entire strata and (b) a weight that accounts for non-response (i.e., lacking test results) among members of our sample. Factor (a) is, crudely, the ratio of number of people in each stratum to the number of people in our sample in the strata; however, the exact weighting accounts for the CPHS and KSS sample design^3^. Factor (b) is a simple ratio of the number of people in our sample to the number of people in our sample for whom we have lab results, without accounting for selection into consent. The percentile confidence intervals for the adjusted proportion per strata are calculated via bootstrap methods with 1000 draws with replacement. Statistical tests are conducted using the percentile confidence intervals.

We estimated the adjusted proportion of positive tests 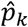 among people in homogenous region *k* as a weighted average of 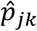 (the adjusted proportion of positive tests in strata (*j, k*)). Our weights for each stratum is the share of the total population in that region that live in that community. The percentile confidence intervals for the adjusted proportion per strata are calculated via bootstrap methods with 1000 draws with replacement. Statistical tests are conducted using the percentile confidence intervals. Similarly, we estimated the adjusted proportion of positive tests 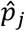 among people in community type *j ∈* urban, rural) as a weighted average of 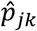 using share of urban/rural population as weights and report bootstrapped confidence intervals for the adjusted proportions.

Finally, we estimate the adjusted seroprevalence using the Rogan-Gladen^4^ correction for imperfect accuracy of tests after calculating adjusted proportions. We employ adjusted proportions to calculate the variance of adjusted seroprevalence and then employ normal approximations to estimate Wald confidence intervals for that prevalence. We employ Microsoft Excel 365 and Stata 16 to perform statistical analyses.

## Results

### Serology tests

The adjusted proportions of positive IgG tests ranged from 22.8-53.1% across rural and 30.9-76.8% across urban areas (Figure 3, details in Table e1). Overall rural, urban and statewide adjusted proportions were 37.4% (95% CI: 32.9-41.8%), 45.6% (95% CI: 38.1-53.1%), and 39.6%, (95% CI: 35.7-43.4%), respectively. Mysore region had a higher adjusted proportion (50.1%, 95% CI: 44.7-55.4%, difference p<0.001). Breakdown of positivity by age is presented in Table 3.

**Table 3.** Seropositivity by Homogeneous Region and Age.

| Homo-<br>genous<br>region | Male | Age Bin | Number of<br>ELISA tests | ELISA<br>positivity<br>rate | Homo-<br>genous<br>region | Male | Age Bin | Number of<br>ELISA tests | ELISA<br>positivity<br>rate |
| --- | --- | --- | --- | --- | --- | --- | --- | --- | --- |
| Bangalore | 1 | 10 | 7 | 0.571429 | Kannada | 0 | 40 | 20 | 0.600000 |
| Bangalore | 0 | 10 | 7 | 0.142857 | Kannada | 1 | 50 | 15 | 0.800000 |
| Bangalore | 1 | 20 | 14 | 0.214286 | Kannada | 0 | 50 | 15 | 0.400000 |
| Bangalore | 0 | 20 | 12 | 0.083333 | Kannada | 1 | 60 | 9 | 0.444444 |
| Bangalore | 1 | 30 | 13 | 0.230769 | Kannada | 0 | 60 | 9 | 0.555556 |
| Bangalore | 0 | 30 | 20 | 0.250000 | Kannada | 1 | 70 | 4 | 0.500000 |
| Bangalore | 1 | 40 | 33 | 0.272727 | Kannada | 0 | 70 | 5 | 0.600000 |
| Bangalore | 0 | 40 | 24 | 0.583333 | Kannada | 1 | 80 | 1 | 1.000000 |
| Bangalore | 1 | 50 | 20 | 0.300000 | Kannada | 0 | 80 | 2 | 1.000000 |
| Bangalore | 0 | 50 | 26 | 0.230769 | Belgaum | 1 | 10 | 8 | 0.375000 |
| Bangalore | 1 | 60 | 26 | 0.461538 | Belgaum | 0 | 10 | 3 | 0.333333 |
| Bangalore | 0 | 60 | 11 | 0.363636 | Belgaum | 1 | 20 | 31 | 0.161290 |
| Bangalore | 1 | 70 | 3 | 0.333333 | Belgaum | 0 | 20 | 16 | 0.375000 |
| Bangalore | 0 | 70 | 4 | 0.250000 | Belgaum | 1 | 30 | 24 | 0.333333 |
| Bangalore | 1 | 80 | 1 | 0.000000 | Belgaum | 0 | 30 | 34 | 0.323529 |
| Bangalore | 0 | 80 | 1 | 0.000000 | Belgaum | 1 | 40 | 69 | 0.376812 |
| Mysore | 1 | 10 | 9 | 0.333333 | Belgaum | 0 | 40 | 31 | 0.322581 |
| Mysore | 0 | 10 | 13 | 0.461538 | Belgaum | 1 | 50 | 51 | 0.333333 |
| Mysore | 1 | 20 | 20 | 0.350000 | Belgaum | 0 | 50 | 25 | 0.320000 |
| Mysore | 0 | 20 | 40 | 0.575000 | Belgaum | 1 | 60 | 10 | 0.500000 |
| Mysore | 1 | 30 | 31 | 0.612903 | Belgaum | 0 | 60 | 7 | 0.285714 |
| Mysore | 0 | 30 | 56 | 0.375000 | Belgaum | 1 | 70 | 2 | 0.500000 |
| Mysore | 1 | 40 | 41 | 0.487805 | Belgaum | 1 | 80 | 1 | 0.000000 |
| Mysore | 0 | 40 | 73 | 0.452055 | Gulbarga | 1 | 10 | 3 | 0.333333 |
| Mysore | 1 | 50 | 27 | 0.444444 | Gulbarga | 0 | 10 | 6 | 0.500000 |
| Mysore | 0 | 50 | 47 | 0.574468 | Gulbarga | 1 | 20 | 11 | 0.272727 |
| Mysore | 1 | 60 | 10 | 0.100000 | Gulbarga | 0 | 20 | 9 | 0.666667 |
| Mysore | 0 | 60 | 8 | 0.375000 | Gulbarga | 1 | 30 | 9 | 0.222222 |
| Mysore | 1 | 70 | 3 | 0.333333 | Gulbarga | 0 | 30 | 13 | 0.230769 |
| Mysore | 0 | 70 | 3 | 0.666667 | Gulbarga | 1 | 40 | 14 | 0.500000 |
| Mysore | 0 | 80 | 2 | 0.500000 | Gulbarga | 0 | 40 | 27 | 0.370370 |
| Kannada | 1 | 10 | 1 | 1.000000 | Gulbarga | 1 | 50 | 16 | 0.437500 |
| Kannada | 0 | 10 | 1 | 0.000000 | Gulbarga | 0 | 50 | 14 | 0.500000 |
| Kannada | 1 | 20 | 8 | 0.625000 | Gulbarga | 1 | 60 | 4 | 0.250000 |
| Kannada | 0 | 20 | 9 | 0.333333 | Gulbarga | 0 | 60 | 6 | 0.500000 |
| Kannada | 1 | 30 | 7 | 0.428571 | Gulbarga | 1 | 70 | 3 | 0.333333 |
| Kannada | 0 | 30 | 14 | 0.571429 | Gulbarga | 0 | 70 | 2 | 0.000000 |
| Kannada | 1 | 40 | 21 | 0.666667 | Gulbarga | 1 | 80 | 1 | 1.000000 |

**Figure 3.**
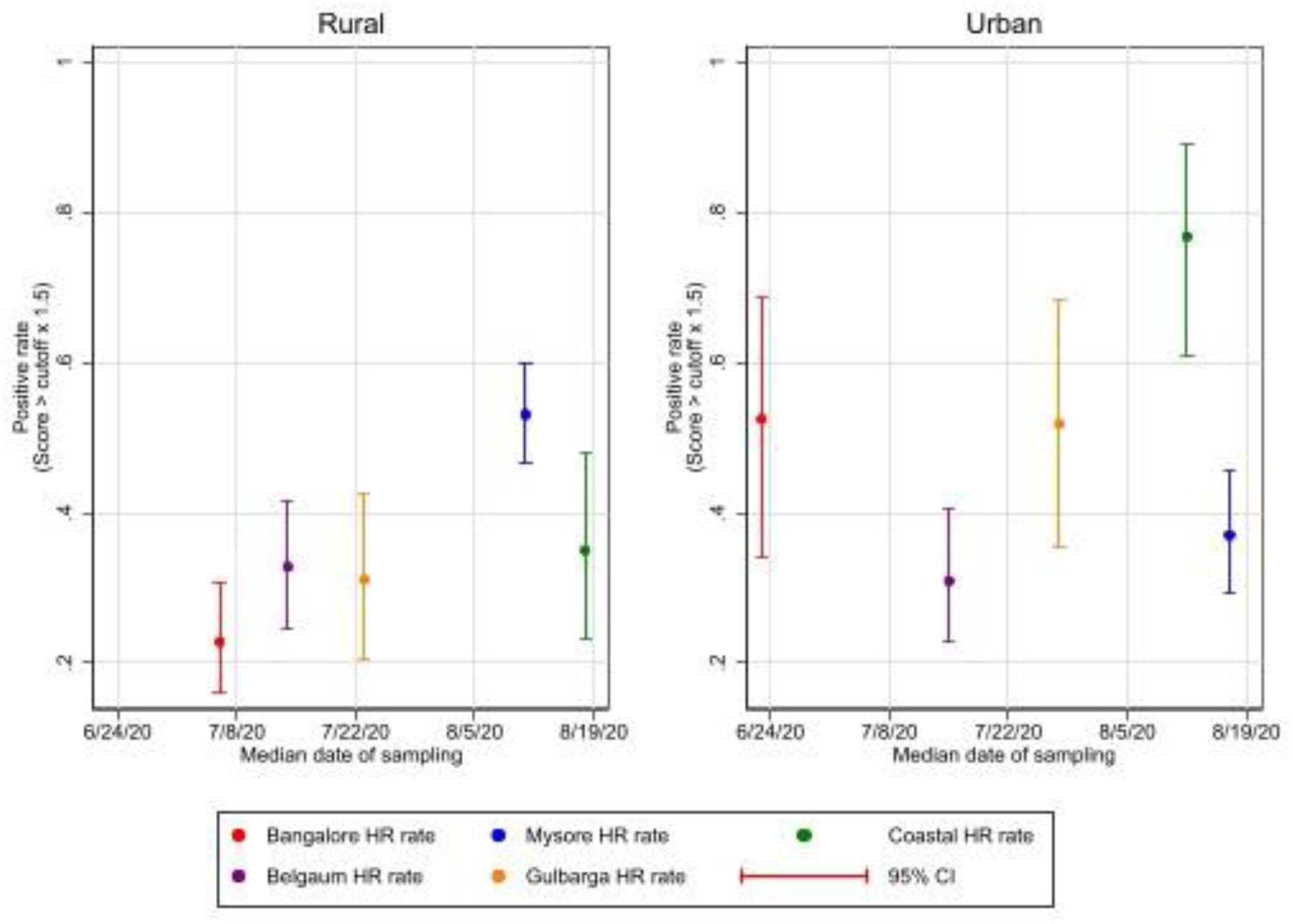
Positive proportion on ELISA-RBD tests, by region and urban status and date. Rural, urban and statewide adjusted-seroprevalences were 44.1 (95% CI: 40.0-48.2%), 53.8 (95% CI: 48.4-59.2%) and 46.7 (95% CI: 43.3-50.0%), respectively.

**Figure 4.**
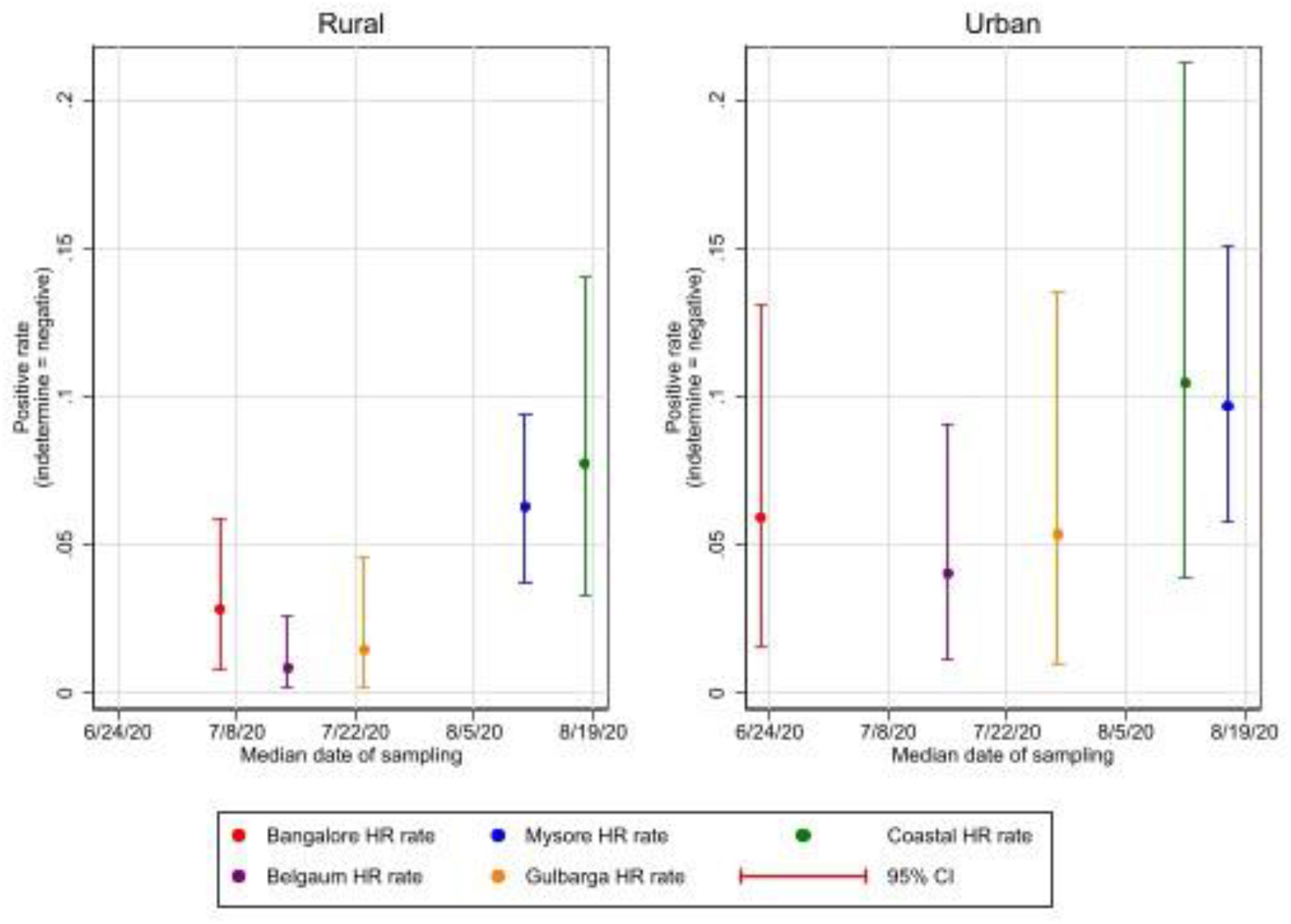
Positive proportion on RT-PCR tests, by region and urban status and by date.

## Discussion

Adjusted seroprevalence across Karnataka implies, based on government mid-year 2020 population estimates, that approximately 31.5 million residents have been infected, 96.4 times the 327,076 publicly reported cases as of August 29, 2020. Estimated RTPCR-positive proportions suggests the epidemic was growing rapidly during August.

Our findings provide new evidence that the COVID-19 epidemic in India has affected rural areas almost as severely as urban areas, despite early attention to the epidemic in urban areas. Two major factors could have contributed to the spread of the epidemic in rural parts of India. First, the release of the lockdown imposed in March was immediately followed by a large migration of daily laborers who lost their sources of income in urban centers and returned home to rural parts of the country. Results from testing large samples of workers to Bihar (a state that received over 2 million returning workers) in May 2020 showed that large share of workers arriving from parts of the country where the epidemic was raging tested positive on RT-PCR^5^. A second contributing factor was that, while urban areas experienced severe lockdowns, rural areas experienced fewer restrictions on mobility (Figure 5) because agricultural activity was deemed an essential sector.

**Figure 5.**
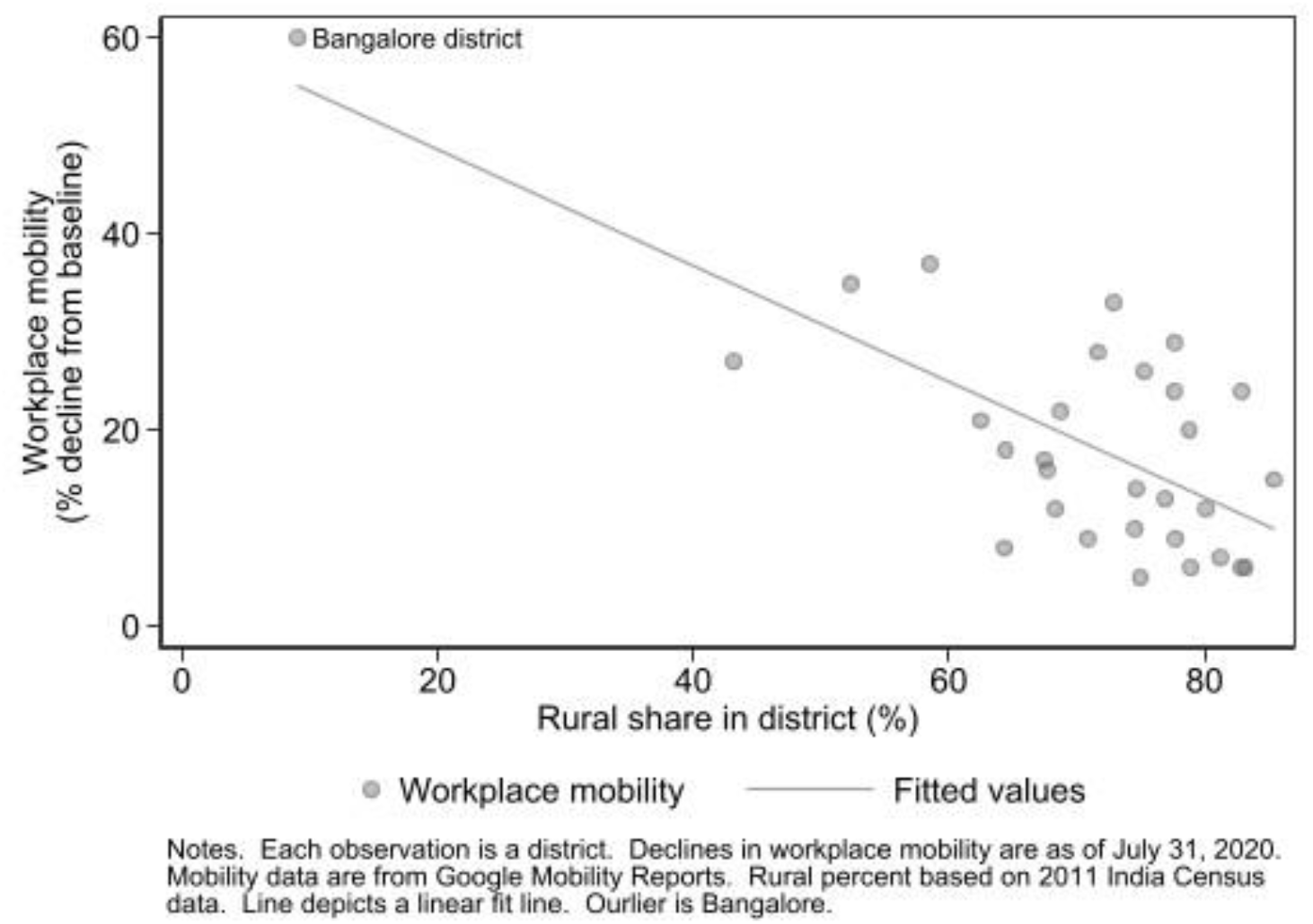
Relationship between rural population share and decline in workplace mobility.

There are several policy implications of our findings. With nearly half the population in the state being infected with COVID as of August 2020, stringent suppression policies across the general population will impose significant costs on those who are already infected. In the short run, most individuals who have already been exposed are likely to be resistant to repeated infection. Until there is further evidence on how fast antibodies (such as those to the RBD of the spike protein we test for) decline over time and whether t-cell immunity provides protection even after antibodies decline, it is difficult to make inferences about long term immunity. However, even in the short run, there is a strong case for adopting frequent testing in exchange for permitting productive economic activity in the state.

That being said, it should be acknowledged that the populations who were exposed in the first half of the epidemic might be significantly different from those who remain uninfected. For example, individuals who are elderly and have chronic conditions or are otherwise at higher risk may have taken precautions to avoid infections. A total relaxation could lead to a spike in infections among such at-risk populations leading to further spikes in severe cases or mortality that will create large burden for the healthcare system. Therefore, as the government considers relaxing restrictions on economic activity, it is critical to continue efforts to promote mask wearing, hand washing, and communicating the significance of COVID complications to individuals who are at risk.

Our findings underscore the need for larger scale studies across India that can provide estimates of seroprevalence at smaller levels of granularity and also study what happens to antibodies and t-cell immunity over time.

## Data Availability

We will make test results available on a de-identified basis 9 months after publication.

## Government and ethical approvals

This study was approved by the Government of India (the Prime Minister’s Principal Scientific Advisor’s Office) and the Government of Karnataka. The study protocols were also approved by IRB / IEC committees at three institutions:

- Karesa (ECR/308/Indt/KA/2018, Approval date June 3, 2020), for Anu Acharya (Mapmygenome)
- Duke University (Protocol 2020-0553),
- University of Chicago (IRB20-1484).

Because Mohanan (Duke) and Malani (University of Chicago) only received de-identified data, the research was determined to be exempt from IRB review at their institutions.

## Acknowledgements

This study was funded by Action Covid-19 Team (ACT) grant awarded to IDFC Foundation (Mumbai, India). We are grateful to the Government of India (K. Vijay Raghavan, Principal Scientific Advisor) and the Government of Karnataka (Jawaid Akhtar, Pankaj Pandey, Dr. Selva Kumar, Gunjan Krishna, Karnataka Covid-19 Technical Advisory Committee, Manoj Kolla, Dr. Prakash, and team members) for supporting and enabling this effort. Thanks to Dr. Gagandeep Kang (THSTI) and Dr. V Ravi (NIMHAS) for timely advice and facilitating the project. We are thankful to Lipika Kapoor and Saloni Taneja for project management and to all team members from CMIE and the project team for undertaking challenging fieldwork during the epidemic. Mr. Manjunath provided outstanding logistics support to enable fieldwork, timely delivery of field supplies and transporting specimens from across the state. All diagnostic testing in the project was conducted at Aster Labs and Xcyton Labs. We thank Reuben Abraham, Pritika Hingorani, and the IDFC Foundation team for supporting all aspects of the project.

## Appendix

**Table e1.** Positive proportion on ELISA (RBD), by homogenous region and area type.

| Region | Cutoff = 1.5 |  |  |  |  |
| --- | --- | --- | --- | --- | --- |
|  | Rural |  |  | Urban |  |
|  | Proportion | (95% CI) |  | Proportion | (95% CI) |
| Bangalore HR | 0.228 | (0.160 0.308) |  | 0.525 | (0.340 0.688) |
| Mysore HR | 0.531 | (0.468 0.598) |  | 0.370 | (0.294 0.456) |
| Coastal HR | 0.350 | (0.231 0.480) |  | 0.768 | (0.610 0.892) |
| Belgaum HR | 0.328 | (0.245 0.415) |  | 0.309 | (0.227 0.405) |
| Gulbarga HR | 0.310 | (0.204 0.425) |  | 0.518 | (0.353 0.685) |

Note. Proportion positive is fraction of tests where the score, defined as IgG response in sample divided by IgG response in known negative sample, is greater than cutoff value. So, e.g., cutoff = 1.5 means test is positive if IgG response in sample is 50% or more greater than in the control sample. Weights are designed to make samples representative of an area (urban or rural) within a region. The weights account for lack of lab results. Bias-corrected bootstrap confidence intervals, based on 1000 replications per estimated proportion, are presented.

**Table e2.** Positive proportion on RT-PCR, by homogenous region and area type.

| Region | Indeterminate coded as negative |  |  |  |  |
| --- | --- | --- | --- | --- | --- |
|  | Rural |  |  | Urban |  |
|  | Proportion | (95% CI) |  | Proportion | (95% CI) |
| Bangalore HR | 0.028 | (0.008 0.058) |  | 0.059 | (0.015 0.131) |
| Mysore HR | 0.063 | (0.037 0.094) |  | 0.097 | (0.058 0.150) |
| Coastal HR | 0.077 | (0.033 0.140) |  | 0.105 | (0.039 0.213) |
| Belgaum HR | 0.008 | (0.002 0.026) |  | 0.040 | (0.011 0.090) |
| Gulbarga HR | 0.015 | (0.002 0.046) |  | 0.054 | (0.009 0.136) |

Note. Proportion positive is fraction of RT-PCR tests where the result is positive. In the top panel, indeterminate results are treated as negative. In the bottom, they are treated as positive. Weights are designed to make samples representative of an area (urban or rural) within a region. The weights account for lack of lab results. Bias-corrected bootstrap confidence intervals, based on 1000 replications per estimated proportion, are presented.

## Footnotes

1 Chaudhuri S, Thiruvengadam R, Chattopadhyay S, et al. Comparative evaluation of SARS-CoV-2 IgG assays in India. Journal of Clinical Virology. 2020;131:104609.

2 See https://www.fda.gov/media/137742/download and https://www.biomerieux-usa.com/sites/subsidiary_us/files/eua-biom-gene-letter.pdf.

3 Details on methods at https://consumerpyramidsdx.cmie.com/kommon/bin/sr.php?kall=wkb.

## References

1. Hallal PC, Hartwig FP, Horta BL, et al. SARS-CoV-2 antibody prevalence in Brazil: results from two successive nationwide serological household surveys. The Lancet Global Health.

2. Chaudhuri S, Thiruvengadam R, Chattopadhyay S, et al. Comparative evaluation of SARS-CoV-2 IgG assays in India. Journal of Clinical Virology. 2020;131:104609.

3. Public Health England. Rapid assessment of Biomerieux ARGENE® SARS-COV-2 R-GENE® real-time detection kit2020.

4. Rogan WJ, Gladen B. Estimating prevalence from the results of a screening test. American journal of epidemiology. 1978;107(1):71–76.

5. Malani A, Mohanan M, Kumar C, Kramer J, Tandel V. Prevalence of SARS-CoV-2 among workers returning to Bihar gives snapshot of COVID across India. medRxiv. 2020:2020.2006.2026.20138545.

